# Hippocampal volumes in UK Biobank are associated with APOE only in older adults

**DOI:** 10.1101/2024.06.05.24307704

**Authors:** Ariya Chaloemtoem, Vera Thornton, Yoonhoo Chang, Andrey P. Anokhin, Michaël E. Belloy, Janine Bijsterbosch, Brian A. Gordon, Sarah M. Hartz, Laura J. Bierut

## Abstract

**INTRODUCTION:** The hippocampus atrophies with age and is implicated in neurodegenerative disorders including Alzheimer’s disease (AD). We examined the interplay between age and *APOE* genotype on total hippocampal volume.

**METHODS:** Utilizing neuroimaging data from 37,463 UK Biobank participants, we applied linear regression to quantify the association of age and *APOE* with hippocampal volume and identified the age when volumes of ε2/ε3, ε3/ε4, and ε4/ε4 carriers significantly deviated from ε3/ε3 using generalized additive modeling.

**RESULTS:** Total hippocampal volume declined with age, with significant differences by *APOE* genotype emerging after age 60. ε3/ε4 and ε4/ε4 carriers displayed reduced volumes from ages 69 and 61, respectively, while ε2/ε3 showed delayed decline starting at age 76.

**DISCUSSION:** The association of *APOE* and hippocampal volume is age-dependent, with differences in volumes of ε4/ε4 carriers detected as early as age 61. This work underscores the importance of *APOE* genotype in determining when to begin screening for AD.

## 1. Introduction

Aging is associated with a gradual decrease in brain volume, a process that accelerates notably in the later stages of life [1]. This decline is evident both globally and regionally across various brain regions. The hippocampus, a pivotal center for cognitive functions such as spatial and episodic memory, is among the most extensively studied regions due to its vulnerability to age-related atrophy and involvement in Alzheimer’s disease (AD) pathogenesis [2]. As one of the earliest and most severely affected regions in AD, the hippocampus exhibits significant neurodegeneration before clinical symptom onset [3].

Increasing age and being a carrier of the apolipoprotein E (*APOE*) ε4 allele are among the greatest risk factors for late onset AD, while *APOE* ε2 is the strongest protective genetic variant [4,5]. The number of ε4 alleles is associated with increased risk of developing AD and earlier age of onset, whereas the number of ε2 alleles is associated with decreased AD risk and later age of onset [5–8]. Some studies have reported that *APOE* ε4 carriers have decreased hippocampal volumes compared to non-carriers, whereas others do not consistently support such associations [9–14]. Similarly, conflicting evidence exists regarding the potential protective role of the ε2 allele in preserving hippocampal volume during aging [9,15,16]. Such discrepancies can be attributed to a range of factors, including differences in study design, sample size, age range of participants, brain imaging protocols, and other potential confounders.

The UK Biobank (UKB) is an excellent resource to address these inconsistencies, offering a large, harmonized neuroimaging dataset with a substantial number of *APOE* ε2 and ε4 allele carriers. The overall UKB cohort comprises over 500,000 community-based participants aged 40 to 69 who were recruited at baseline assessment between 2006 and 2010 [17]. Since 2014, a subset of participants returned for neuroimaging assessments and data are currently available for approximately 45,000 participants [18]. Leveraging this large, population-based neuroimaging sample of participants with *APOE* genotypes ε3/ε3, ε3/ε4, ε4/ε4, and ε2/ε3, our goal is to examine the association of *APOE* and total hippocampal volume across age and identify the age at which divergence from the ε3/ε3 status becomes detectable.

## 2. Methods

### 2.1 Participants

Our study leveraged data from the UKB, an extensive research initiative conducted in the United Kingdom. Details of the UKB resource are publicly available [17]. UKB obtained ethical approval from the North West Centre for Research Ethics Committee (Ref: 11/NW/0382). All participants provided written informed at baseline assessments and later at imaging assessments. This study was conducted under UKB Research Application Numbers 47267 and 48123. All participants were included regardless of self-reported race or ethnicity, or genetic ancestry.

Of 42,801 participants with neuroimaging data available as of January 25, 2023, 37,463 were retained in our study. We first filtered for individuals with available *APOE* genotype and neuroimaging data, as well as active consent as of April 25, 2023. Individuals with neurological diseases were then removed (Supplementary Table 1) and then filtered such that all remaining participants were genetically not third-degree or closer relatives (Supplementary Text). See Supplementary Figure 1 for a graphical summary and Supplementary Table 2 for the distribution of *APOE* genotypes by sample filtering step. Only those with *APOE* ε3/ε3, ε3/ε4, ε4/ε4, or ε2/ε3 were included in our analysis. The age range of the participants spanned from 44 to 82 with a mean of 63.9 years (SD = 7.7).

### 2.2 Brain imaging phenotypes

Magnetic resonance imaging (MRI) brain images were acquired on Siemen’s Skyra 3T scanners with a 32-channel head coil (Siemen’s Medical Solutions, Germany) across multiple UKB sites. The acquired imaging data were processed using standardized pipelines developed by the UKB imaging team and made available to researchers as summary statistics representing key brain imaging variables referred to as imaging derived phenotypes (IDPs). Full details of the image acquisition protocol and processing pipeline are publicly available [19,20].

We utilized UKB data on left and right hippocampal volumes generated through subcortical volumetric sub-segmentation using FreeSurfer (N = 41,238). Specifically, we were interested in examining the combined total hippocampal volume obtained by summing the left and right measures (Data-field IDs: 26641 and 26663, respectively).

### 2.3 *APOE* genotype

UKB provides genotypes obtained from two closely related genotyping arrays: the UK BiLEVE Axiom Array and the UKB Axiom Array and additional variants imputed using the Haplotype Reference Consortium and the UK10K + 1000 Genomes reference panels. Full genotyping and quality control details are described by Bycroft and are publicly available [21]. Using the phased haplotype data supplied by UKB, we derived the two *APOE* alleles for each participant, based on single nucleotide polymorphism (SNP) markers rs429358 and rs7412 on chromosome 19. The *APOE* genotype of each participant was then assigned based on the combination of the two alleles. The resulting *APOE* allele and genotype frequencies before and after sample filtering are shown in Supplementary Tables 3 and 4.

### 2.4 Covariates

Several covariates were included to account for potential confounding factors. These include the site of imaging, as well as scanning date, head size, and resting state functional MRI motion, which was further split by site, as recommended by UKB [22]. We also incorporated covariates related to participant demographics and lifestyle commonly reported as risk factors for AD. These include genetic sex, body mass index (BMI), years of education, and income, as well as lifetime history of daily smoking, lifetime history of pack years smoked, drinking status, and past year alcohol consumption (drinks per week) [23]. To account for population structure, we included the first ten genetic principal components (PCs) computed by UKB using principal component analysis (PCA). Total gray matter volume, which decreases with age, was also added as a covariate to allow us to assess differences in hippocampal volume relative to the rest of the brain.

The UKB data-fields corresponding to each variable are outlined in Supplementary Table 5. Covariates were obtained from the imaging assessment and missing values were backfilled with baseline survey responses (Supplementary Table 6). The multiple imputation by chained equations (MICE) method with the classification and regression trees (CART) approach was then used to impute any remaining missing values [24]. Detailed explanation of variable processing handling of missing data can be found in the Supplementary Text.

### 2.5 Statistical analysis

Chi-square tests and analysis of variance (ANOVA) were applied to examine the distribution and underlying patterns of demographic and lifestyle variables by *APOE* genotype.

We then examined the trajectory of total hippocampal volume across age by *APOE* genotype using a model-free sliding-window approach, which does not assume a linear relationship between the variables. We used a sliding interval of 1 year and window length of ±5 years. Mean volumes were calculated within each window and plotted as a function of age along with 95 percent confidence intervals. This procedure was performed separately on raw hippocampal volume and residual volume after regressing out covariate effects.

To formally assess the interaction between age and *APOE* status, we fit two linear regression models:

Model 1 for the additive effects of *APOE* and age:

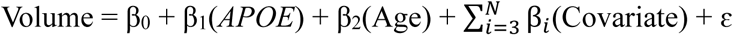

Model 2 for the interactive effects *APOE* and age:

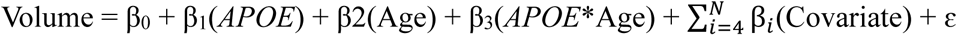

Full model equations are provided in the Supplementary Text. *APOE* genotype was treated as a categorical variable with four levels: ε2/ε3, ε3/ε3, ε3/ε4, and ε4/ε4. The ε3/ε3 genotype served as the reference group for comparison. Age was split into three groups (<60, 60-69, and 70+) based on sliding-window results. The youngest age group (<60 years) served as the reference category for the remaining groups (60-69 and 70+). Categorizing age allows us to account for its non-linear association with hippocampal volume, while categorizing *APOE* by genotype avoids assuming an additive linear effect of ε2 or ε4 allele dosage. This approach also facilitates the interpretation of results. See Supplementary Table 7 for details on the data type and variable encoding of each predictor. The goodness-of-fit between the two models was assessed using a likelihood ratio test (LRT).

Next, to determine the age at which total hippocampal volumes of those with ε2/ε3, ε3/ε4, and ε4/ε4 diverge from that of ε3/ε3 carriers, we modeled its association with age for each *APOE* genotype using a generalized additive model (GAM) with a cubic regression spline using the ‘mgcv’ R package (version 1.9) [25]. Pairwise differences between fitted smooths were then estimated and the point at which ε2/ε3, ε3/ε4, and ε4/ε4 diverged from ε3/ε3 was defined as the age when the confidence intervals no longer overlapped.

All statistical analyses were conducted using R statistical software (version 4.1.2), and a significance level of α = .05 was used to determine statistical significance. Our code is available at https://github.com/achaloemtoem/ukb_hippocampus.

## 3. Results

### 3.1 Sample characteristics

Our final sample consisted of 37,463 participants, with an average age of 63.9 years (SD = 7.7). Of those, 13% had *APOE* ε2/ε3, 61% had *APOE* ε3/ε3, 24% had *APOE* ε3/ε4, and 2% had *APOE* ε4/ε4. Table 1 shows the study sample characteristics, including mean age with standard deviation, as well as the number of participants in each age group, by *APOE* genotype. There was a significant difference in mean age and the distribution of age categories by *APOE* genotype (P < .001), reflecting a smaller proportion of older participants with ε3/ε4 and ε4/ε4 in our data (see Supplementary Figure 2). This finding is consistent with previously reported differential survival based on *APOE* status [26]. Aside from BMI (P < .01), all other sociodemographic and lifestyle characteristics did not differ among *APOE* groups.

**Table 1.** Demographics and sample characteristics by *APOE* genotype.

|  | Total | *APOE* genotype |  |  |  | *P* |
| --- | --- | --- | --- | --- | --- | --- |
|  |  | ε2/ε3 | ε3/ε3 | ε3/ε4 | ε4/ε4 |  |
| **N** | 37,463 | 4,826 | 22,720 | 9,048 | 869 |  |
| **Age (years)** | 63.9 ± 7.7 | 64.1 ± 7.7 | 64.0 ± 7.7 | 63.5 ± 7.6 | 63.3 ± 7.4 | <.001 |
| **Age group, n (%)** |  |  |  |  |  |  |
| <60 | 11,497 (31) | 1,439 (30) | 6,848 (30) | 2,925 (32) | 285 (33) | <.001* |
| 60-69 | 15,969 (42) | 2,035 (42) | 9,663 (43) | 3,880 (43) | 391 (45) |  |
| ≥70 | 9,997 (27) | 1,352 (28) | 6,209 (27) | 2,243 (25) | 193 (22) |  |
| **Sex, n (%)** |  |  |  |  |  |  |
| Female | 1,9819 (53) | 2,549 (53) | 11,919 (52) | 4,871 (54) | 480 (55) | ns* |
| Male | 1,7644 (47) | 2,277 (47) | 10,801 (48) | 4,177 (46) | 389 (45) |  |
| **BMI (kg/m<sup>2</sup>)** | 26.5 ± 4.4 | 26.6 ± 4.4 | 26.5 ± 4.3 | 26.4 ± 4.4 | 26.0 ± 4.1 | <.01 |
| **Education (years)** | 15.9 ± 4.7 | 15.8 ± 4.8 | 15.9 ± 4.7 | 15.9 ± 4.7 | 16.0 ± 4.8 | ns |
| **Income (£), n (%)** |  |  |  |  |  |  |
| <18,000 | 4,413 (12) | 580 (13) | 2,630 (12) | 1,096 (13) | 107 (13) | ns* |
| 18,000-30,999 | 9,681 (27) | 1,308 (28) | 5,908 (27) | 2,236 (26) | 229 (27) |  |
| 31,000-51,999 | 10,868 (30) | 1,378 (30) | 6,534 (30) | 2,713 (31) | 243 (29) |  |
| 52,000-100,000 | 8,353 (23) | 1,036 (22) | 5,084 (23) | 2,040 (23) | 193 (23) |  |
| ≥100,000 | 2,644 (7) | 331 (7) | 1,605 (7) | 638 (7) | 70 (8) |  |
| **Ever smoked daily, n (%)** |  |  |  |  |  |  |
| Yes | 9,291 (25) | 1,233 (26) | 5,583 (25) | 2,277 (25) | 198 (23) | ns* |
| No | 28,165 (75) | 3,592 (74) | 17,133 (75) | 6,769 (75) | 671 (77) |  |
| **Pack years of smoking** | 4.7 ± 11.2 | 4.9 ± 11.7 | 4.7 ± 11.2 | 4.6 ± 10.8 | 4.0 ± 10.7 | ns |
| **Drinking status, n (%)** |  |  |  |  |  |  |
| Current | 35,042 (94) | 4,532 (94) | 21,268 (94) | 8,430 (93) | 812 (93) | ns* |
| Former | 1,217 (3) | 159 (3) | 727 (3) | 307 (3) | 24 (3) |  |
| Never | 1,203 (3) | 135 (3) | 724 (3) | 311 (3) | 33 (4) |  |
| **Drinks per week** | 7.6 ± 8.4 | 7.5 ± 8.0 | 7.6 ± 8.4 | 7.6 ± 8.4 | 7.6 ± 8.9 | ns |
| **Hippocampal volume (cm<sup>3</sup>)** | 7.5 ± 0.8 | 7.4 ± 0.8 | 7.5 ± 0.8 | 7.5 ± 0.8 | 7.4 ± 0.8 | <.001 |
| **Total gray matter volume (cm<sup>3</sup>)** | 664.9 ± 59.5 | 663.1 ± 59.6 | 665.1 ± 59.4 | 665.3 ± 59.7 | 664.8 ± 57.8 | ns |

Values are reported as mean ± standard deviation. Reported *P* values reflect ANOVA test results. ns, not significant
\* *P* value represents results of $\chi^2$ test

### 3.2 Effect of age and *APOE* on hippocampal volume

Hippocampal volume decreased with age across all *APOE* genotypes. This decline was evident when examining the moving averages of hippocampal volume as well as when age was categorized into three groups (Figure 1 and Supplementary Figures 3 and 4). The raw total hippocampal volumes by *APOE* and age group are summarized in Supplementary Table 8.

**Figure 1.**
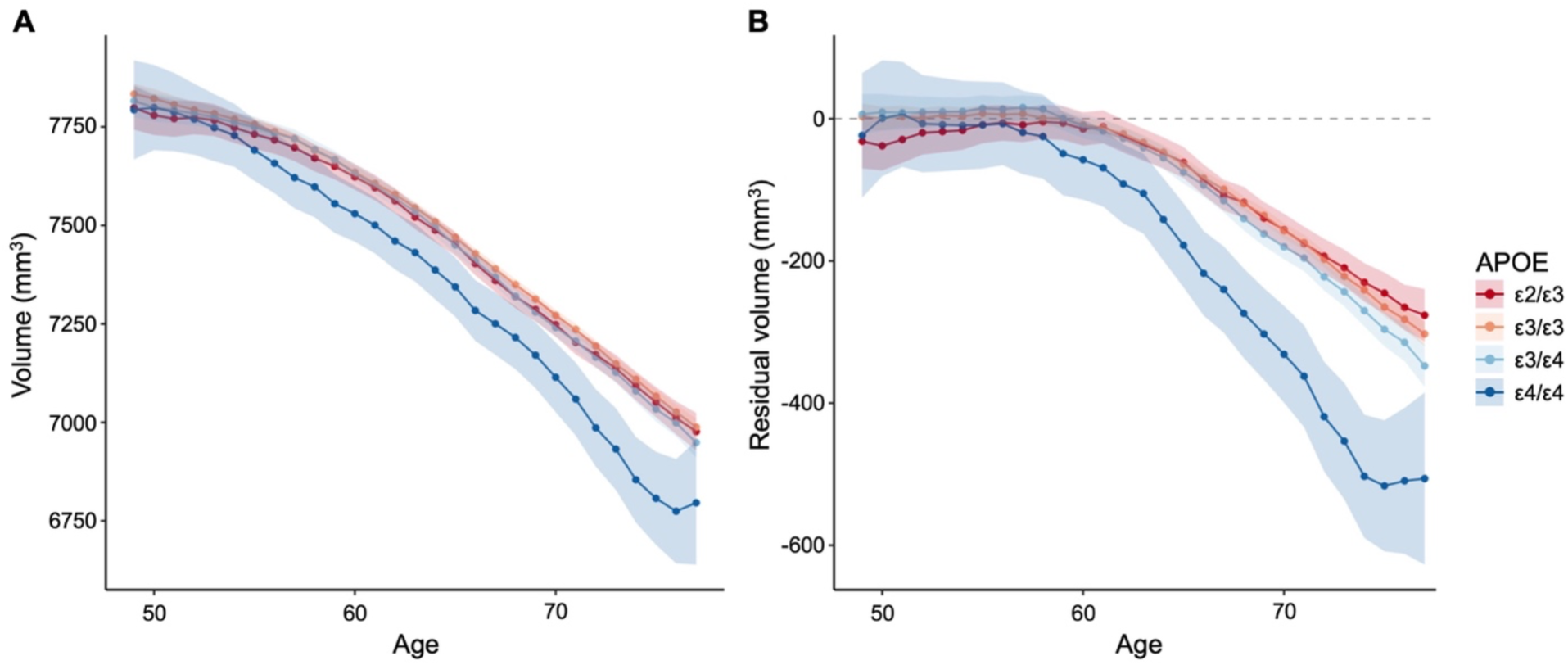
5-year centered moving average of (A) raw total hippocampal volume and (B) residual total hippocampal volume after adjusting for covariates and centering at the mean of ε3/ε3 age <60.

We then fit two regression models to explore the main and interactive effects of age and *APOE* status on total hippocampal volume with categorical age (Table 2). In our first model examining the main effects in the absence of interactions between age and *APOE*, we observed a robust negative and non-linear relationship between total hippocampal volume and age. Specifically, participants in the 60 to 69 age group had a 116.1 mm^3^ reduction in hippocampal volume compared to those younger than 60 (P = 7.4×10^-61^), and this effect magnified over threefold in individuals aged 70 and older (β = −407.0 mm^3^, P < 10^-300^).

**Table 2.** Regression results for total hippocampal volume (mm^3^)

|  | Model 1* |  | Model 2† |  |
| --- | --- | --- | --- | --- |
|  | Beta | P | Beta | P |
| ***APOE* effect** |  |  |  |  |
| ε2/ε3 | -2.5 | 0.77 | -18.2 | 0.24 |
| ε3/ε3 (ref) | - | - | - | - |
| ε3/ε4 | -11.7 | 0.08 | 5.4 | 0.65 |
| ε4/ε4 | -109.1 | 3.9e<sup>-9</sup> | -10.1 | 0.76 |
| **Age effect** |  |  |  |  |
| <60 (ref) | - | - | - | - |
| 60-69 | -116.1 | 7.4e<sup>-61</sup> | -111.3 | 2.7e<sup>-36</sup> |
| ≥70 | -407.0 | <1e<sup>-300</sup> | -397.0 | 3.3e<sup>-298</sup> |
| ***APOE* × Age effect** |  |  |  |  |
| ε2/ε3 60-69 | - | - | 13.2 | 0.51 |
| ≥70 | - | - | 36.3 | 0.10 |
| ε3/ε4 60-69 | - | - | -15.9 | 0.31 |
| ≥70 | - | - | -40.3 | 2.3e<sup>-2</sup> |
| ε4/ε4 60-69 | - | - | -97.6 | 2.2e<sup>-2</sup> |
| ≥70 | - | - | -246.2 | 1.3e<sup>-6</sup> |

Model comparison based on the LRT, P = 1.1e<sup>-5</sup>
\* Model 1: Volume = $\beta_0 + \beta_1(APOE) + \beta_2(Age) + \sum_{i=3}^N \beta_i(Covariate) + \varepsilon$
† Model 2: Volume = $\beta_0 + \beta_1(APOE) + \beta_2(Age) + \beta_3(APOE*Age) + \sum_{i=4}^N \beta_i(Covariate) + \varepsilon$

In addition, we observed a main effect of *APOE* status on hippocampal volume only in ε4/ε4 carriers. Individuals with the ε4/ε4 genotype displayed a reduction in total hippocampal volume of 109.1 mm^3^ compared to ε3/ε3 carriers (P = P = 3.9×10^-9^). We did not detect statistically significant associations of ε3/ε4 or ε2/ε3 and total hippocampal volume in this model.

Adding interaction terms between age and *APOE* to our model significantly improved its fit (P = 1.1×10^-5^). A robust negative and non-linear association between total hippocampal volume and age remains with similar effect size estimates compared to the model without the interaction terms. However, we no longer observe a main effect of *APOE* status on hippocampal volume. Instead, we found significant interactions between age and *APOE* genotype (Figure 2). Among participants under 60 years old, *APOE* had no effect on hippocampal volume (Table 2). The age-dependent effect of *APOE* became apparent in older participants. Specifically, among those aged 60 to 69, homozygosity for the ε4 allele was significantly associated with hippocampal volume loss (β = −97.6 mm^3^, P = .022), and the largest age-dependent effect of *APOE* was observed in individuals aged 70 and older carrying two copies of the ε4 allele (β = −246.2 mm^3^, P = 1.3×10^-6^). We found no significant interactive effect of ε3/ε4 among those aged 60 to 69, but in the 70 and older age group we did observe significant associations of *APOE* ε3/ε4 with decreased hippocampal volume (β = −40.3 mm^3^, P = .023). No interactions were found in ε2/ε3 carriers.

**Figure 2.**
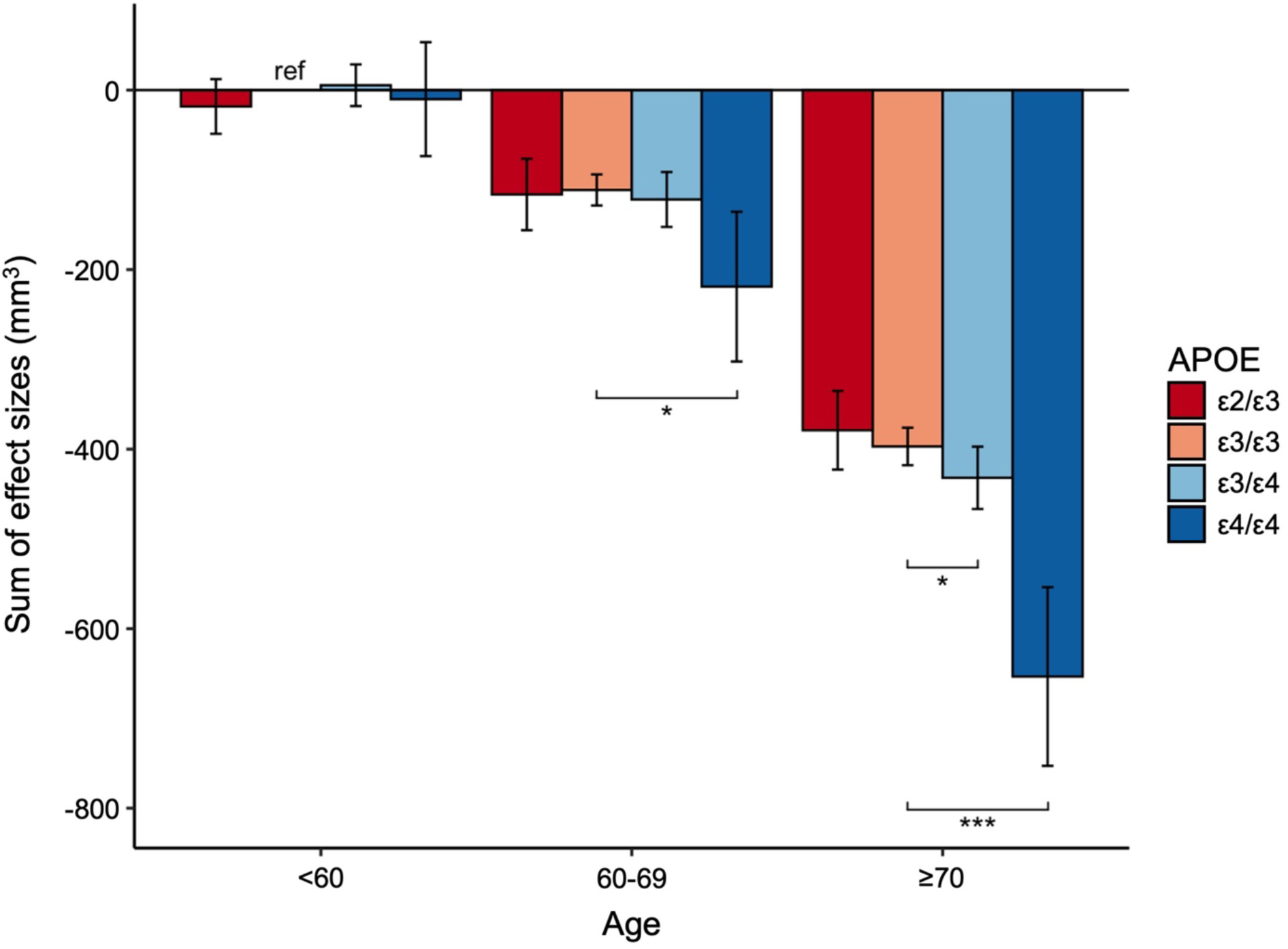
Sum of effect sizes for total hippocampal volume (mm^3^) relative to age <60 and ε3/ε3 status based on results from the regression model with interactions. Error bars represent 95% CI and asterisks indicate significant interactions based on linear regression results. * P < 0.05, ** P < 0.01, *** P < 0.001

We further examined whether *APOE* and age showed comparable associations with total grey matter volume as they did with total hippocampal volume. We found that total grey matter volume decreased with age, but there was no effect of *APOE* nor evidence of an interactive effect between *APOE* and age (Supplementary Tables 9 and 10, and Supplementary Figures 5 and 6). This finding supports the notion that the interactive effect of *APOE* genotype we observed is specific to hippocampal volume loss and is independent of overall brain atrophy associated with aging.

### 3.3 Cross-sectional age of divergence by *APOE* genotype

We used GAM to analyze the relationship between hippocampal volume and age, stratified by *APOE* genotype. Our aim was to identify the age at which volumes of ε2/ε3, ε3/ε4, and ε4/ε4 carriers significantly diverged from those of ε3/ε3 carriers (Figure 3). Our analysis confirmed a significant non-linear relationship between hippocampal volume and age across *APOE* genotypes (P < 2.0×10^-16^), as expected. Hippocampal volume in ε4/ε4 carriers began to deviate significantly from the ε3/ε3 trajectory at age 61, showing an accelerated decline, while significant deviation for ε3/ε4 carriers occurred at age 69. Conversely, ε2/ε3 carriers show a slower rate of decline in hippocampal volume compared to ε3/ε3 carriers starting at age 76.

**Figure 3.**
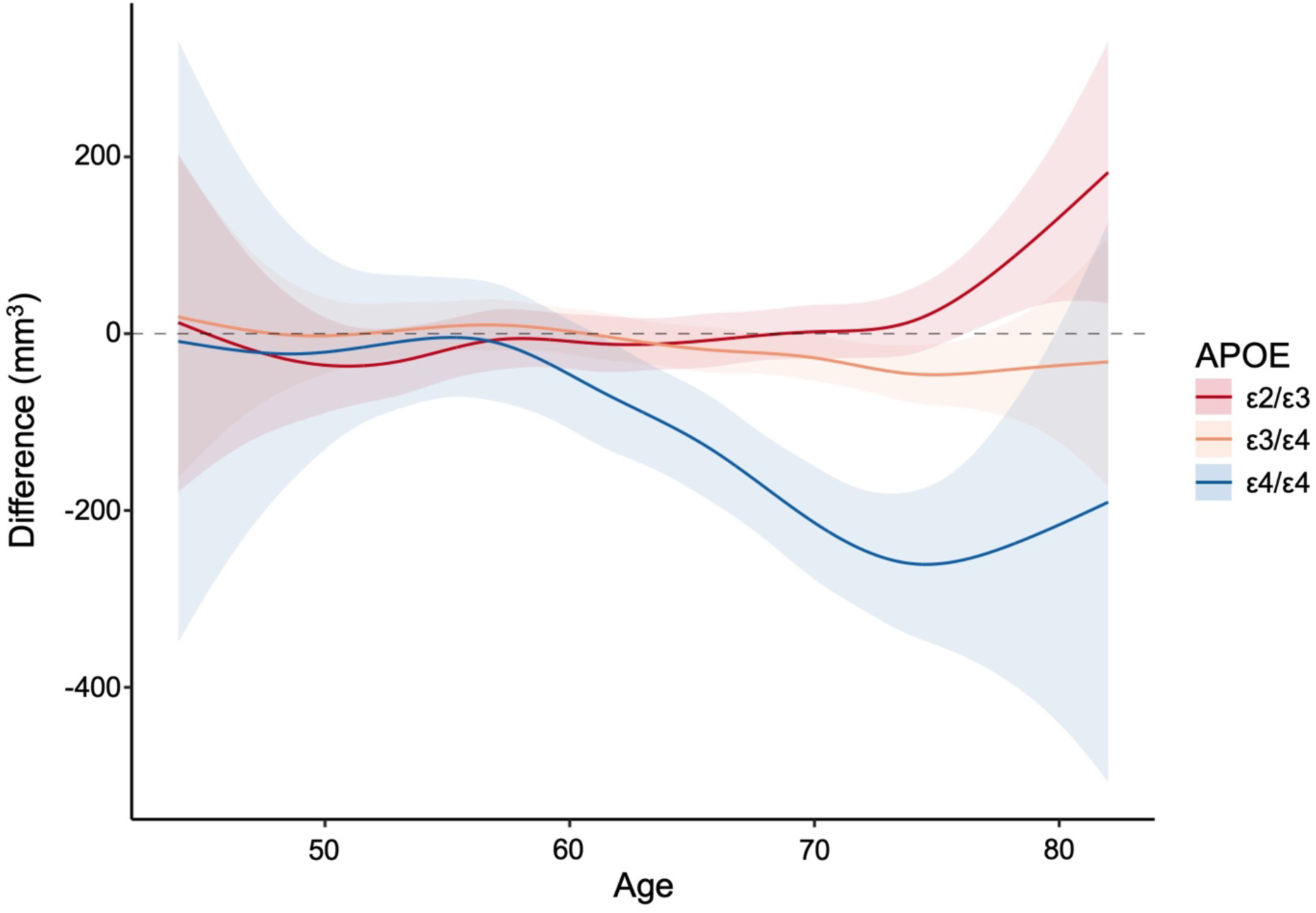
Estimated differences in total hippocampal volume from ε3/ε3 by *APOE* genotype.

## 4. Discussion

We found significant age-dependent associations of *APOE* and total hippocampal volume in a large, cross-sectional, community-based sample of adults. Prior to the age of 60, *APOE* genotype had no discernible effects on total hippocampal volume. Significant modifying effects of ε3/ε4 and ε4/ε4 were found only in older participants. Specifically, the earliest age of divergence from the trajectory of hippocampal volume loss in ε3/ε3 carriers was detected at age 61 for ε4/ε4 carriers and 69 for ε3/ε4 carriers, demonstrating accelerated volume loss. Although we did not find a significant interaction between *APOE* ε2/ε3 and age, our findings suggest a potential role of *APOE* ε2/ε3 in mitigating hippocampal volume loss around age 76. These results underscore the complex interplay between *APOE* genotype and age in shaping hippocampal volume trajectories.

The association between *APOE* and measures of hippocampal atrophy has been investigated in numerous studies, but discrepancies in findings persist. These inconsistencies have raised concerns about the statistical power and reproducibility of neuroimaging studies, many of which feature small sample sizes [27–29]. Contributing to this variability are differences in study designs, such as comparisons of AD patients or cognitively impaired individuals to non-affected controls, as well as variations in the treatment and coding of the *APOE* variable. In many cases, due to the low frequency of ε4 homozygotes, researchers have combined heterozygotes (ε3/ε4) and homozygotes (ε4/ε4) for comparison with ε3/ε3 carriers [30–32]. However, grouping ε3/ε4 and ε4/ε4 genotypes masks differences in their effects across the lifespan. Others have focused their analysis on ɛ4 gene-dose effects, typically defined by pooling participants according to the number of ε4 alleles and treating *APOE* genotype as continuous numeric variable, which assumes a linear additive effect of the allele [9,14].

Our study takes advantage of the large sample size of the UKB neuroimaging cohort to build upon existing knowledge by examining the effects of categorized *APOE* genotypes in community-based individuals without dementia. In a recent UKB-based cross-sectional study, Veldsman et al. (2020) analyzed hippocampal volume trends across age and *APOE*, and our findings are consistent with their results [33]. In addition, they concentrated solely on ε3/ε4 and ε4/ε4 carriers given established reductions in hippocampal volume. In our analysis we include ε2/ε3 heterozygotes. This broader approach is warranted given the large sample of ε2/ε3 carriers in the UKB and previously reported opposing effects of *APOE* ε4 and ε2 alleles on hippocampal morphology [9].

It is crucial to note that our findings are contingent upon detectable genotype differences in our sample which may be limited in the older age range. Although the UKB neuroimaging cohort features a large number of ε4/ε4 carriers (N = 869), we observe a decreasing proportion of ε4 carriers with advancing age (Supplementary Figure 2A). This trend aligns with previous studies reporting lower ε4 frequencies with increasing age, partly due to earlier mortality and lower survival rates [26]. These dynamics impact our ability to accurately capture the effects of the ε4 allele in the older age groups, particularly those over age 70.

Another limitation of our study lies in its cross-sectional nature, which constrains our ability to assess individual-level rates of change over time. However, despite this limitation, our findings align with those reported by Mishra et al. (2018) in a longitudinal study involving 497 cognitively normal middle to older age participants [34]. They categorized participants as either *APOE* ε4 carriers (ε3/ε4 or ε4/ε4) or non-carriers (ε2/ε2, ε2/ε3, or ε3/ε3) and observed greater rate of hippocampal atrophy in *APOE* ε4 carriers relative to non-carriers, with this effect first detected at age 57.

Our findings diverge from those of Walhovd et al. (2020) who studied 1,181 cognitively healthy participants from age 4 to 95 years under the hypothesis that AD genetic risk factors exert their effects throughout the lifespan rather than only at later life stages [35]. The authors categorized participants as either *APOE* ε4 carriers or non-carriers and found that *APOE* ε4 was associated with lower hippocampal volumes early in life, creating an initial offset. However, they found no significant interactions between *APOE* and age that would indicate faster hippocampal atrophy in older age. In contrast, we find no differences in hippocampal volume in those under the age of 60, and we provide compelling evidence for such interactive effects. This is noteworthy given that structural MRI indicators of neurodegeneration is considered an advanced stage biomarker of AD [36,37].

Recent advancements in AD research have shown that commercially available blood-based biomarkers can identify AD pathology with high accuracy and prior to the onset of cognitive symptoms [38]. Our work identifies when structural MRI changes in hippocampal volume associated with *APOE* genotype, a later disease stage biomarker, can be detected. These findings suggest that biomarker testing should begin earlier for ε3/ε4 and ε4/ε4 carriers compared to ε3/ε3 carriers. Integrating information from these different modalities could improve screening, diagnostic accuracy and disease monitoring, facilitating timely and personalized prevention and treatment interventions.

In summary, this study identified no differences in hippocampal volume by *APOE* status in those under the age of 60 and then accelerated hippocampal volume loss in ε4/ε4 carriers compared to ε3/ε3 carriers starting at age 61, and in ε3/ε4 carriers at age 69. Conversely, we observed reduced volume loss in ε2/ε3 carriers beginning at age 76. These findings underscore the critical role of age in modifying the impact of the *APOE* status on brain structure and carries significant clinical implications for screening and potential prevention of AD.

## Data Availability

All data produced in the present work are contained in the manuscript.

## Acknowledgements

This work was supported by the National Institutes of Health (NIH) National Institute on Alcohol Abuse and Alcoholism (NIAAA) grant numbers U10AA008401 [AC, APA, LJB, and YC] and R01AA029308 [AC and SMH]. LJB is additionally funded by NIAAA grant number R01AA027049. SMH is further supported by the National Institute on Aging (NIA) grant R01AG065234. BAG is funded by NIA grants P01AG026276, U19AG032438, R01AG065234, R01AG073267, R01AG079569, R01AG070139, R01AG073424 and R01AG082030, as well as the National Institute of Child Health and Human Development (NICHD) grant R21HD112910. APA is funded by NIAAA grant number R01025646 and the National Institute on Drug Abuse (NIDA) grant number R01DA058114. JB is funded by the National Institute of Mental Health (NIMH) grants R01MH128286 and R01MH132962. MEB is supported by NIA grant R00AG075238. VT is additionally funded by Washington University Institute of Clinical and Translational Sciences grant number TL1TR002344.

The funding sources of this study had no role in the study design, the collection, the analysis or the interpretation of data, in the writing of the report, or in the decision to submit the article for publication. This research was conducted using the UK Biobank Resource under Application Numbers 47267 and 48123. We extend our sincere appreciation to the participants who generously dedicated their time to contribute to the UK Biobank study.

## Consent Statement

The UK Biobank received ethical approval from the North West Centre for Research Ethics Committee (Ref: 11/NW/0382). All participants provided signed informed written consent prior to participation.

## Abbreviations

AD: Alzheimer’s disease
*APOE*: apolipoprotein E
GAM: generalized additive model
IDP: imaging derived phenotype
MRI: magnetic resonance imaging
UKB: UK Biobank
SD: standard deviation

