## Supplemental Material for "Hippocampal volumes in UK Biobank are associated with APOE only in older adults"

*Chaloemtoem et al.*

**Supplementary Text**

Removing participants with neurological diagnoses

Participants with pre-existing neurological conditions are removed from our sample. During both the baseline and imaging appointments participants were asked about their medical history and all diagnoses were recorded using codes in data-field 20002. We removed all participants with a neurological condition reported in either visit that might potentially affect their imaging based on a list described by Gray et al [1]. Supplementary Table 1 summarizes the list of neurological conditions, their corresponding data codes, and the number of participants affected in our sample.

Removing 3rd degree or closer relatives

Related individuals in the UKB tend to cluster geographically due to the influence of family participation and the location of assessment centers [2]. We excluded all 3rd degree or closer relatives in our final sample. Relatives of 3rd degree or closer were identified using a kinship matrix provided by UKB. We first filtered for related pairs where both individuals are in the neuroimaging set and identified clusters of related pairs. Within those clusters, we implemented a modified version of an algorithm developed by Hanscombe et al [3] to drop participants in the order of the most highly interconnected until only pairs of related individuals remain. We added a condition to prioritize retaining rarer *APOE* genotypes (ε4/ε4 > ε3/ε4 > ε2/ε3 > ε3/ε3). In cases where the most highly interconnected pairs within a cluster shared the same *APOE* genotype, we randomly selected one member of the pair for removal. The code is available here.

Deriving the alcohol and smoking measures

UKB participants were first asked to indicate alcohol use frequency (Data-field 1558: Alcohol intake frequency). For those reporting current drinking, participants estimated their typical weekly intake (for daily to weekly drinkers) or monthly intake (for those drinking less frequently) in units of red wine, white wine, fortified wine, beer and cider, spirits, or other (such as alcopops). If intake varied, participants were instructed to estimate their average intake over the past year. To derive a standard measure, we summed drinks across different drink types to get the total units of drinks consumed per week or per month. For participants providing a monthly estimate, this total was divided by 4.3 to obtain a weekly estimate. We then used this information combined with data-field 3731 (denoting participants who formerly drank alcohol), which was asked only of those who indicated "never" in data-field 1558 (Alcohol intake frequency), to distinguish between those who never drank and those who formerly drank and assign a drinking status ("current", "former", or "never") to each participant. Current drinking status was assigned to those with a non-zero weekly estimated intake whereas the never status was assigned to those with a weekly estimated past year alcohol intake of 0 who also did not formerly drink. The code is available here.

For smoking measures, participants indicated current (Data-field 1239: Current tobacco smoking) or past smoking (Data-field 1249: Past tobacco smoking). For those reporting daily or near-daily smoking, which constituted approximately 25% of the sample, additional questions were asked about the age of smoking onset and quantity of cigarettes smoked to compute pack years (Data-field 20161). Approximately half of the sample reported either never smoking or smoking fewer than 100 times in their lifetime. For these participants, 0 pack years were assigned. Those endorsing occasional smoking with more than 100 cigarettes over their lifetime ("yes" in Data-field 2644) were assigned 0.5 pack years. The code is available here.

We applied this method of derivation separately to survey responses collected during the baseline and imaging visits.

Deriving covariates other than alcohol and smoking

Covariates that required further processing or recoding prior to backfilling and missing data imputation include years of education, income, and select imaging covariates recommended by UKB [4]. The imaging covariates we processed according to UKB recommendations are site of imaging, scanning date, head size, and resting state functional MRI (rfMRI) motion. We describe how each of these covariates were processed below and code is available here.

Years of education

Data-field 6138 asks participants about their educational qualifications (College of University degree, A levels/AS levels or equivalent, O levels/GCSEs or equivalent, CSEs or equivalent, NVQ or HND or HNC or equivalent, Other professional qualifications e.g. nursing, teaching). We converted these qualifications into years of education using the method employed by Zhou et al [5], which maps each qualification to an International Standard Classification of Education category and imputes the equivalent years of education for each category.

Income

Income, as recorded in data-field 738, is categorical, with participants selecting their income band from predefined ranges (less than 18000, 18000 to 30999, 31000 to 52000, 52000 to 100000, and greater than 100000). We converted these categories into a continuous numeric value by using the lower bound of each income band.

Imaging covariates

We processed our imaging covariates in accordance with the procedure described by Alfaro-Almagro et al [4]. The date of attending the imaging site was converted into a numeric format using the as.numeric() function in R, which transforms the date into the number of days since January 1, 1970. All imaging covariates, excluding imaging site were demeaned and normalized using the median and median absolute deviation * 1.48 (equivalent to one standard deviation). Outliers greater than 8 were removed and replaced the symbol NA (not available), denoting a missing value. Given that site is considered the most influential covariate [4], the date, head size, and rfMRI variables were split on site. Within each site, missing values and outliers were replaced with the site median. The values for each site (except the site value itself) were then z-score normalized within that site. We included $date^2$ to account for non-linear effects of date. Finally, the per-site confounds were renamed to create unique variables for each site in the format site#_variable (e.g., site1_date), where these variables retain the values for their corresponding site and zeros for the other two sites.

Supplementary Table 7 presents an overview of the data types for each covariate, as well as for all other variables incorporated in our analyses that were not described in this section.

Handling of missing data

For all covariates, we prioritize data from the imaging visit as the primary phenotypic variable. In cases where this data is missing, we substitute it with the corresponding value from the baseline survey, if available. Responses such as “Do not know” and “Prefer not to answer” are replaced with NA and considered as missing data. Any remaining missing values after backfilling (see Supplementary Table 6) are imputed using the multiple imputation by chained equations (MICE) method with the classification and regression trees (CART) approach (code available here). The covariates with missing values are imputed based on the observed values of the other variables in an interactive process, adjusting for any correlations or dependencies between the variables in the dataset. Variables that did not have any missing values before performing imputation using MICE include age, sex, and BMI.

Full regression equations
Detailed equations of the regression models used for testing the additive and interactive effects of *APOE* and age on hippocampal volume are described below:

Model 1 for the additive effects of *APOE* and age:

```
Hippocampal volume ~ e2e3 + e3e4 + e4e4 + age group (60-69) + age group (70+) +
                sex + BMI + years of education + income + total gray matter volume +
                ever daily smoked + pack years + drinking status + drinks per week +
                site + site1_date + site1_date^2 + site1_head size + site1_rfMRI motion +
                site2_date + site2_date^2 + site2_head size + site2_rfMRI motion +
                site3_date + site3_date^2 + site3_head size + site3_rfMRI motion +
                PC_1 + PC_2 + PC_3 + PC_4 + PC_5 + PC_6 + PC_7 + PC_8 + PC_9 + PC_10
```

Model 2 for the interactive effects of *APOE* and age:

```
Hippocampal volume ~ e2e3 + e3e4 + e4e4 + age group (60-69) + age group (70+) +
                e2e3:age group (60-69) + e3e4:age group (60-69) + e4e4:age group (60-69) +
                e2e3:age group (70+) + e3e4:age group (70+) + e4e4:age group (70+)  +
                sex + BMI + years of education + income + total gray matter volume +
                ever daily smoked + pack years + drinking status + drinks per week +
                site + site1_date + site1_date^2 + site1_head size + site1_rfMRI motion +
                site2_date + site2_date^2 + site2_head size + site2_rfMRI motion +
                site3_date + site3_date^2 + site3_head size + site3_rfMRI motion +
                PC_1 + PC_2 + PC_3 + PC_4 + PC_5 + PC_6 + PC_7 + PC_8 + PC_9 + PC_10
```

We examined the additive and interactive effects of *APOE* and age on total gray matter volume using the same equation as for hippocampal volume, but without including total gray matter volume as a predictor variable. Variable encodings are outlined in Supplementary Table 7.

**Supplementary Table 1.** Neurological condition diagnosis codes* (N = 1,597)

| Condition | Code | N |
|---|---|---|
| Stroke or ischemic stroke | 1081 | 539 |
| Epilepsy | 1264 | 351 |
| Transient ischemic stroke | 1082 | 325 |
| Meningitis | 1247 | 258 |
| Multiple sclerosis | 1261 | 204 |
| Head injury | 1266 | 154 |
| Parkinson's | 1262 | 87 |
| Subarachnoid hemorrhage | 1086 | 37 |
| Encephalitis | 1246 | 30 |
| Guillain-Barre syndrome | 1256 | 28 |
| Brain hemorrhage | 1491 | 20 |
| Dementia/Alzheimer's/cognitive impairment | 1263 | 15 |
| Meningioma | 1659 | 14 |
| Ischemic stroke | 1583 | 13 |
| Neurological disease/trauma | 1240 | 13 |
| Cerebral aneurysm | 1425 | 10 |
| Motor neuron disease | 1259 | 10 |
| Spina bifida | 1524 | 10 |
| Subdural hematoma | 1083 | 10 |
| Other demyelinating disease | 1397 | 9 |
| Chronic degenerative neurological | 1258 | 8 |
| Brain/intracranial abscess | 1245 | 5 |
| Cerebral palsy | 1433 | 4 |
| Nervous system infection | 1244 | 1 |

* Diagnosis codes are from data-field 20002. Participants may have more than one diagnosis.

**Supplementary Table 2.** N by sample filtering step and *APOE* genotype after selecting for those with FreeSurfer-based hippocampal volume measures

| | | *APOE* genotype | | | | | |
|---|---|---|---|---|---|---|---|
| **Step** | **Total** | **ε2/ε2** | **ε2/ε3** | **ε2/ε4** | **ε3/ε3** | **ε3/ε4** | **ε4/ε4** |
| Import | 41,148 | 234 | 5,124 | 962 | 24,385 | 9,534 | 909 |
| Consent withdrawn removed | 41,147 | 234 | 5,124 | 962 | 24,385 | 9,533 | 909 |
| Neurological diagnosis removed | 39,550 | 223 | 4,929 | 917 | 23,445 | 9,162 | 874 |
| Related (≥$3^{rd}$ degree) removed | 38,555 | 214 | 4,826 | 878 | 22,720 | 9,048 | 869 |

**Supplementary Table 3.** *APOE* allele frequencies before and after sample filtering

| | **Before** (N = 41,148) | | **After** (N = 38,555) | |
|---|---|---|---|---|
| **Allele** | N alleles | % | N alleles | % |
| ε2 | 6,554 | 8.0 | 6,132 | 8.0 |
| ε3 | 63,418 | 77.1 | 59,314 | 76.9 |
| ε4 | 12,314 | 14.9 | 11,664 | 15.1 |

**Supplementary Table 4.** *APOE* genotype frequencies before and after sample filtering

| | **Before** (N = 41,148) | | **After** (N = 38,555) | |
|---|---|---|---|---|
| **Genotype** | N genotypes | % | N genotypes | % |
| ε2/ε2 | 234 | 0.6 | 214 | 0.6 |
| ε2/ε3 | 5,124 | 12.5 | 4,826 | 12.5 |
| ε2/ε4 | 962 | 2.3 | 878 | 2.3 |
| ε3/ε3 | 24,385 | 59.3 | 22,720 | 58.9 |
| ε3/ε4 | 9,534 | 23.2 | 9,048 | 23.5 |
| ε4/ε4 | 909 | 2.2 | 869 | 2.3 |

**Supplementary Table 5.** Variables and corresponding UK Biobank data-field ID(s)

| Variable | Data-field ID |
|---|---|
| Age | 21003 |
| Genetic sex | 22001 |
| Body Mass Index (BMI) | 21001 |
| Education | 6138 |
| Income | 738 |
| Smoking | |
| Ever daily smoked* | 1239 (current), 1249 (past) |
| Pack years of smoking† | 20161 (pack years), 2644 (light smokers) |
| Alcohol | |
| Drinking status‡ | Drinks per week§ and 3731 (former) |
| Drinks per week§ | 1558 (alcohol intake frequency)<br>Monthly dosage: 4407, 4418, 4429, 4440, 4451, 4462<br>Weekly dosage: 1568, 1578, 1588, 1598, 1608, 5364 |
| Date attended assessment center | 53 |
| Imaging site | 54 |
| Head size | 25000 |
| Hippocampal volume | 26641 (left), 26715 (right) |
| Total gray matter volume | 26518 |
| rfMRI motion | 25741 |
| Neurological diagnosis codes | 20002 |

* Denotes lifetime history of daily smoking created from touchscreen questionnaire on current and past smoking
† Assigned to those with a history of daily smoking as defined by UKB which we modified to also assign values to light smokers who have smoked at least 100 cigarettes in their lifetime
‡ Derived from drinks per week variable combined with touchscreen questionnaire on former drinking
§ Created from touchscreen questionnaire on past year dose of different types of alcohol

**Supplementary Table 6.** Summary of missing data before and after backfilling for covariates that were backfilled (N = 38,555)

| Covariate | Before N | Before % | After N | After % |
|---|---|---|---|---|
| BMI | 1,313 | 3.41 | 0 | 0.00 |
| Drinking status | 268 | 0.70 | 1 | 0.00 |
| Drinks per week | 272 | 0.71 | 33 | 0.09 |
| Education | 386 | 1.00 | 18 | 0.05 |
| Ever daily smoked | 370 | 0.96 | 7 | 0.02 |
| Income | 3,848 | 9.98 | 1,550 | 4.02 |
| Pack years of smoking | 1,357 | 3.52 | 417 | 1.08 |

**Supplementary Table 7.** Data type of each variable

| Variable | Type | Details |
|---|---|---|
| *APOE* | Categorical | 4 levels: ε2/ε3, ε3/ε3, ε3/ε4, and ε4/ε4<br>(dummy coded with ε3/ε3 as the reference genotype) |
| Age | Categorical | 3 levels: <60, 60-69 and 70+<br>(dummy coded with <60 as the reference age group) |
| Sex | Binary | 0 = Female<br>1 = Male |
| BMI | Continuous | - |
| Education | Numeric | Number of years of education as described in the supplementary text |
| Income | Numeric | See supplementary text |
| Ever daily smoked | Binary | 0 = No, denoting no history of daily smoking<br>1 = Yes, indicating a history of daily smoking |
| Pack years of smoking | Continuous | See supplementary text |
| Drinking status | Categorical | 3 levels: Never, Former, and Current |
| Drinks per week | Continuous | See supplementary text |
| Site | Categorical | 3 levels: Site 1, Site 2, and Site 3 |
| Date | Continuous | See supplementary text |
| Head size | Continuous | See supplementary text |
| rfMRI motion | Continuous | See supplementary text |

**Supplementary Table 8.** Raw total hippocampal volume ($cm^3$) by age and *APOE* (N = 37,463)

| | ε2/ε3 | | ε3/ε3 | | ε3/ε4 | | ε4/ε4 | |
|---|---|---|---|---|---|---|---|---|
| **Age group** | Mean | 95% CI | Mean | 95% CI | Mean | 95% CI | Mean | 95% CI |
| <60 | 7.75 | [7.71, 7.78] | 7.77 | [7.75, 7.79] | 7.76 | [7.74, 7.79] | 7.73 | [7.65, 7.81] |
| 60-69 | 7.48 | [7.44, 7.51] | 7.50 | [7.48, 7.51] | 7.48 | [7.46, 7.50] | 7.37 | [7.29, 7.45] |
| 70+ | 7.05 | [7.01, 7.09] | 7.06 | [7.05, 7.08] | 7.03 | [7.00, 7.06] | 6.81 | [6.69, 6.93] |

CI = confidence interval

**Supplementary Table 9.** Regression results for total gray matter volume ($mm^3$)

| | Model 1 | | Model 2 | |
|---|---|---|---|---|
| | Beta | P | Beta | P |
| ***APOE* effect** | | | | |
| ε2/ε3 | 73 | 0.87 | 97 | 0.91 |
| ε3/ε3 (ref) | - | - | - | - |
| ε3/ε4 | -141 | 0.68 | -391 | 0.53 |
| ε4/ε4 | -295 | 0.76 | -904 | 0.59 |
| **Age effect** | | | | |
| <60 (ref) | - | - | - | - |
| 60-69 | -18699 | ~0 | -18743 | ~0 |
| ≥70 | -39526 | ~0 | -39571 | ~0 |
| ***APOE* × Age effect** | | | | |
| ε2/ε3 60-69 | - | - | -332 | 0.75 |
| ≥70 | - | - | 417 | 0.72 |
| ε3/ε4 60-69 | - | - | 485 | 0.55 |
| ≥70 | - | - | 166 | 0.86 |
| ε4/ε4 60-69 | - | - | -1378 | 0.54 |
| ≥70 | - | - | -2605 | 0.33 |

**Supplementary Table 10.** Raw total gray matter volume($cm^3$) by age and *APOE* (N = 37,463)

| | ε2/ε3 | | ε3/ε3 | | ε3/ε4 | | ε4/ε4 | |
|---|---|---|---|---|---|---|---|---|
| **Age group** | Mean | 95% CI | Mean | 95% CI | Mean | 95% CI | Mean | 95% CI |
| <60 | 682 | [679, 685] | 682 | [681, 684] | 681 | [679, 683] | 679 | [673, 686] |
| 60-69 | 662 | [660, 665] | 664 | [663, 665] | 663 | [662, 665] | 663 | [657, 668] |
| 70+ | 644 | [641, 647] | 648 | [647, 649] | 648 | [646, 651] | 648 | [640, 657] |

CI = confidence interval

**Supplementary Figure 1.** Flow chart of sample filtering. Asterisk denotes too few participants imaged at a fourth site. They were excluded to minimize batch effects.

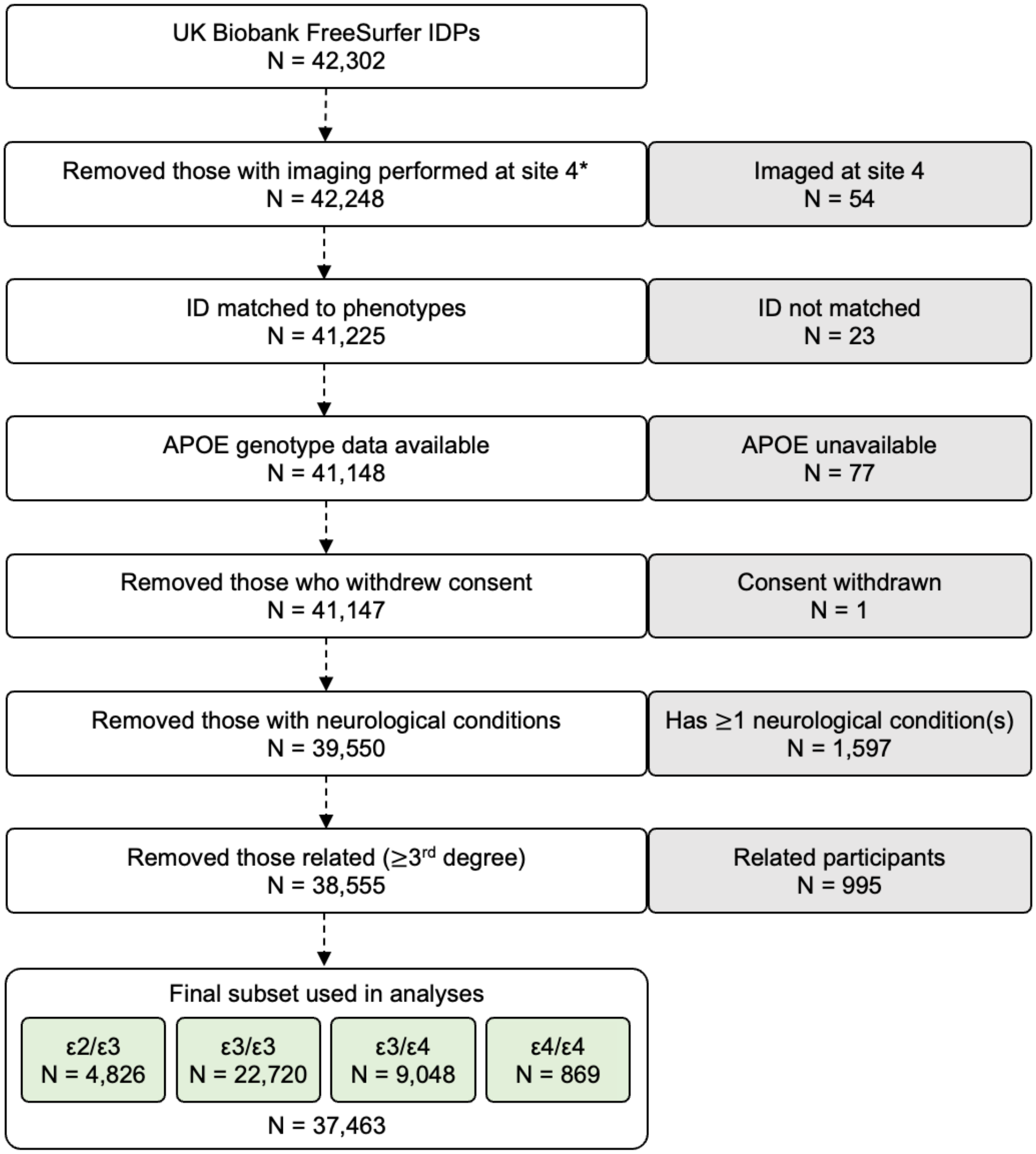

**Supplementary Figure 2.** Distribution of *APOE* (A) allele and (B) genotype frequencies

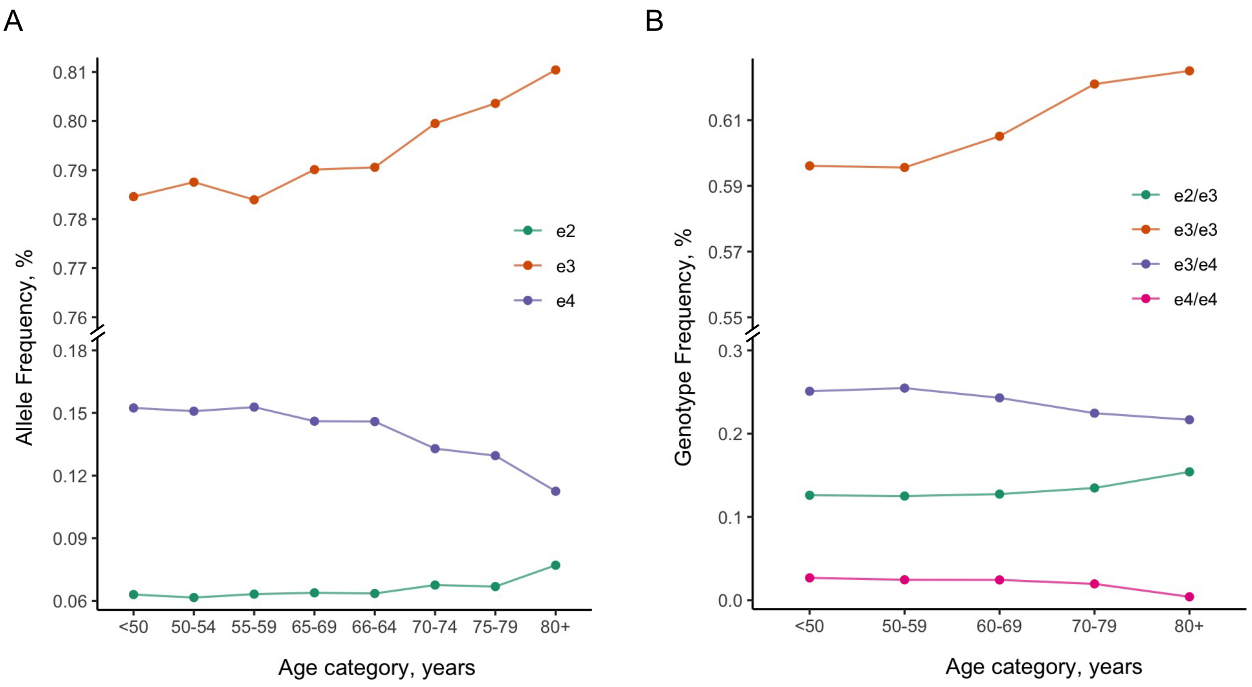

**Supplementary Figure 3.** Distribution of raw hippocampal volume by (A) age and (B) *APOE*, Distribution of residual hippocampal volume by (C) age and (D) *APOE* ($mm^3$). Asterisks indicate significant pairwise t-test results. Bonferroni adjusted P *< .05, **< .01, ***< .001, ****< .001 (N = 37,463)

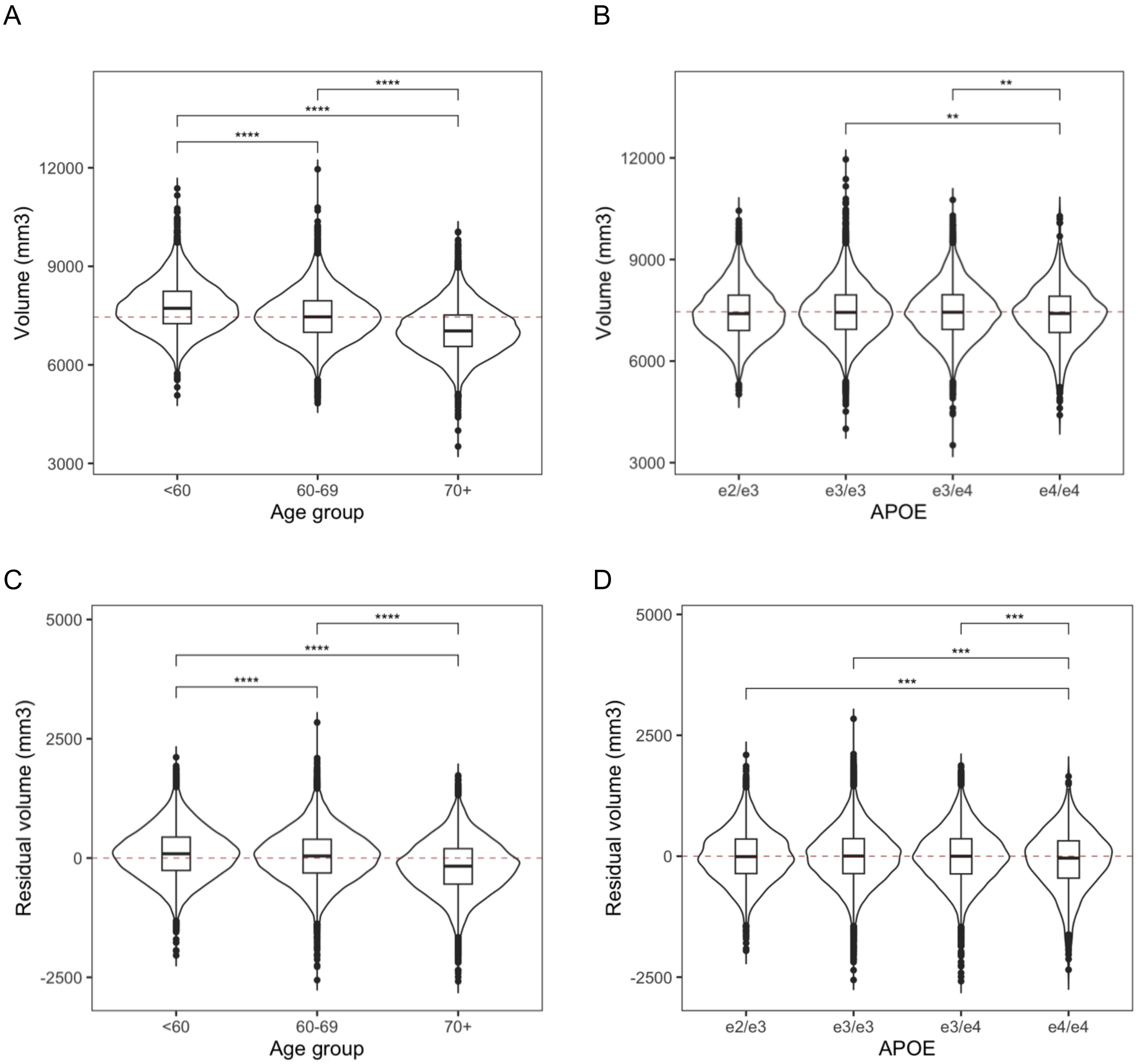

**Supplementary Figure 4.** Group-level average of (A) raw hippocampal volume and (B) residual hippocampal volume after adjusting for covariates and centering at ε3/ε3 age <60 ($mm^3$) (N = 37,463)

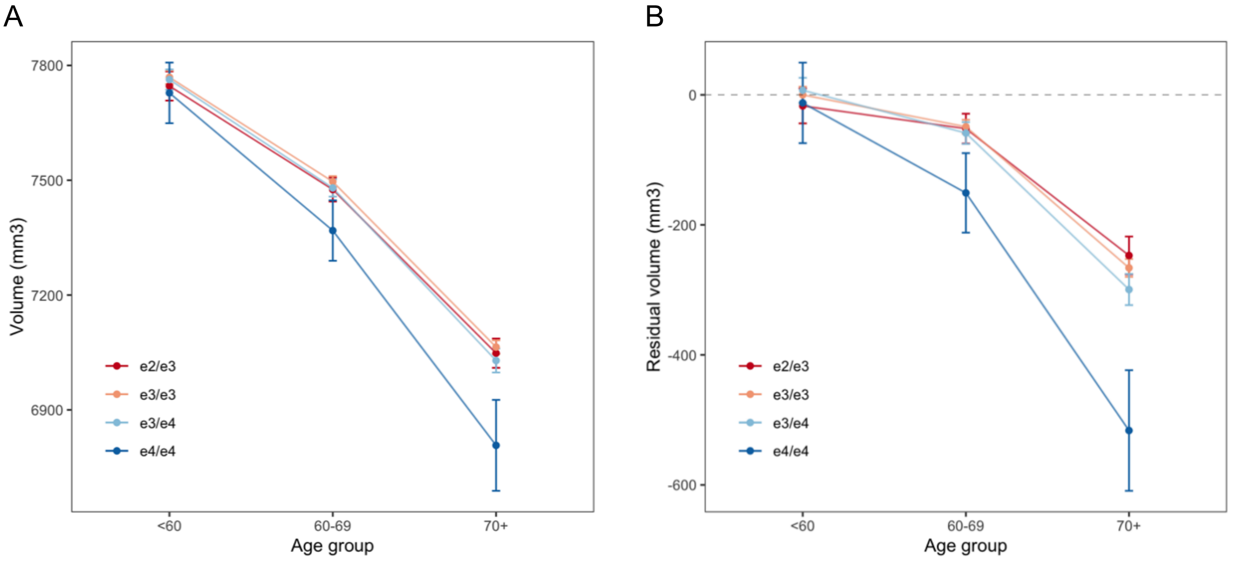

**Supplementary Figure 5.** Sum of effect sizes for total gray matter volume ($mm^3$) relative to age <60 and ε3/ε3. Error bars represent 95% confidence intervals. (N = 37,463)

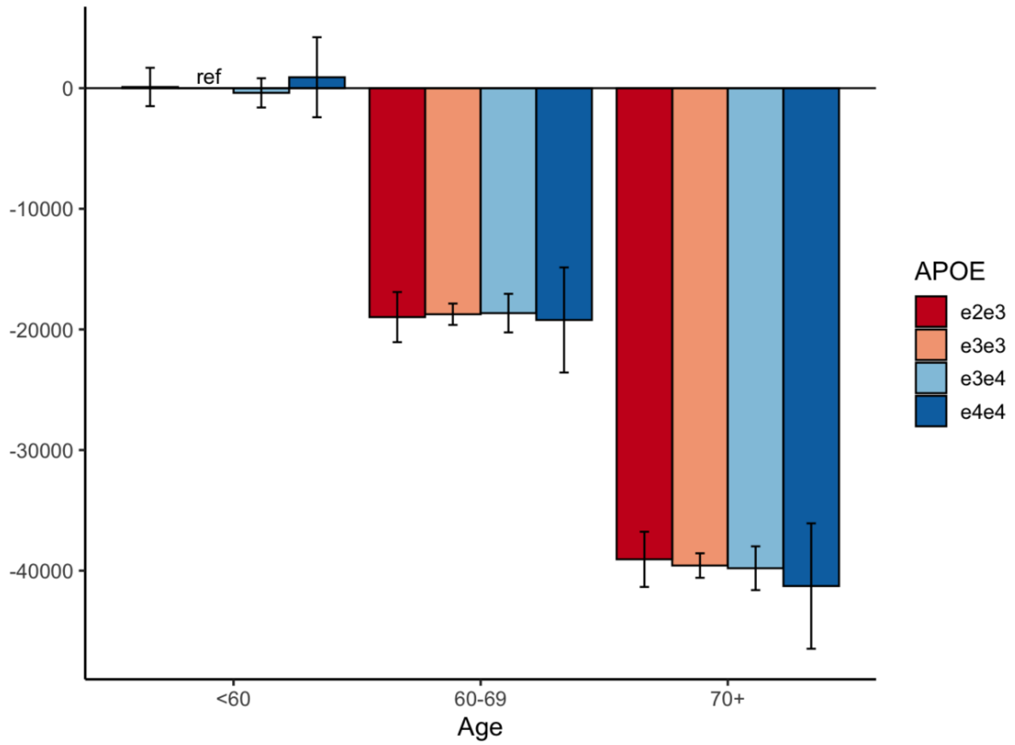

**Supplementary Figure 6.** Distribution of raw total gray matter volume by (A) age and (B) *APOE*, Distribution of residual total gray matter volume by (C) age and (D) *APOE* ($mm^3$). Asterisks indicate significant pairwise t-test results. Bonferroni adjusted P *< .05, **< .01, ***< .001, ****< .001 (N = 37,463)

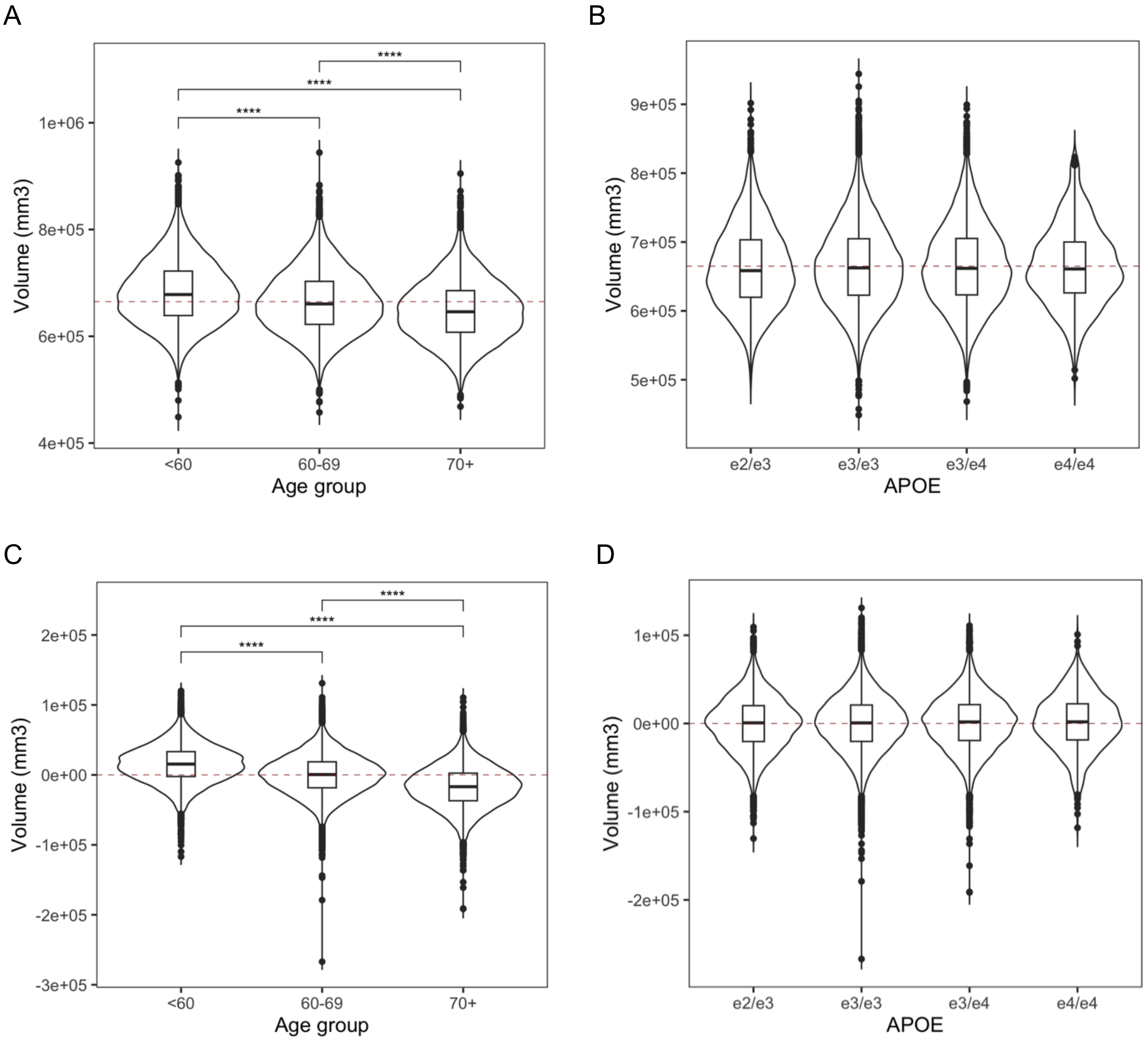
